# Chymase in Plasma and Urine Extracellular Vesicles: Novel Biomarkers for Primary Hypertension

**DOI:** 10.1101/2023.11.09.23298324

**Authors:** Sarfaraz Ahmad, Gagan Deep, Henry A Punzi, Yixin Su, Sangeeta Singh, Ashish Kumar, Shalini Mishra, Amit K Saha, Kendra N Wright, Jessica L VonCannon, Louis J Dell’Italia, Wayne J Meredith, Carlos M Ferrario

## Abstract

**BACKGROUND:** Extracellular vesicles (EVs) have emerged as a promising liquid biopsy for various diseases. For the first time, using plasma and urinary EVs, we assessed the activity of renin-angiotensin system (RAS), a central regulator of renal, cardiac, and vascular physiology, in patients with control (Group I) or uncontrolled (Group II) primary hypertension.

**METHODS:** EVs were isolated from 34 patients with history of hypertension, and characterized for size and concentration by nanoparticle tracking analyses, exosomal biomarkers by immunogold labeling coupled with transmission electron microscopy, flow cytometry and immunoblotting. EVs were analyzed for the hydrolytic activity of chymase, angiotensin converting enzyme (ACE), ACE2, and neprilysin (NEP) by HPLC.

**RESULTS:** Plasma and urinary EVs were enriched for small EVs and expressed exosomal markers (CD63, CD9, and CD81). The size of urinary EVs (but not plasma EVs) was significantly larger in Group II compared to Group I. Differential activity of RAS enzymes was observed, with significantly higher chymase activity compared to ACE, ACE2, and NEP in plasma EVs. Similarly, urinary EVs exhibited higher chymase and NEP activity compared to ACE and ACE2 activity. Importantly, compared to Group I, significantly higher chymase activity was observed in urinary EVs (p = 0.03) from Group II, while no significant difference in activity was observed for other RAS enzymes.

**CONCLUSIONS:** Bioactive RAS enzymes are present in plasma and urinary EVs. Detecting chymase in plasma and urinary EVs uncovers a novel mechanism of angiotensin II-forming enzyme and could also mediate cell-cell communication and modulate signaling pathways in recipient cells.

**GRAPHICAL ABSTRACT:** 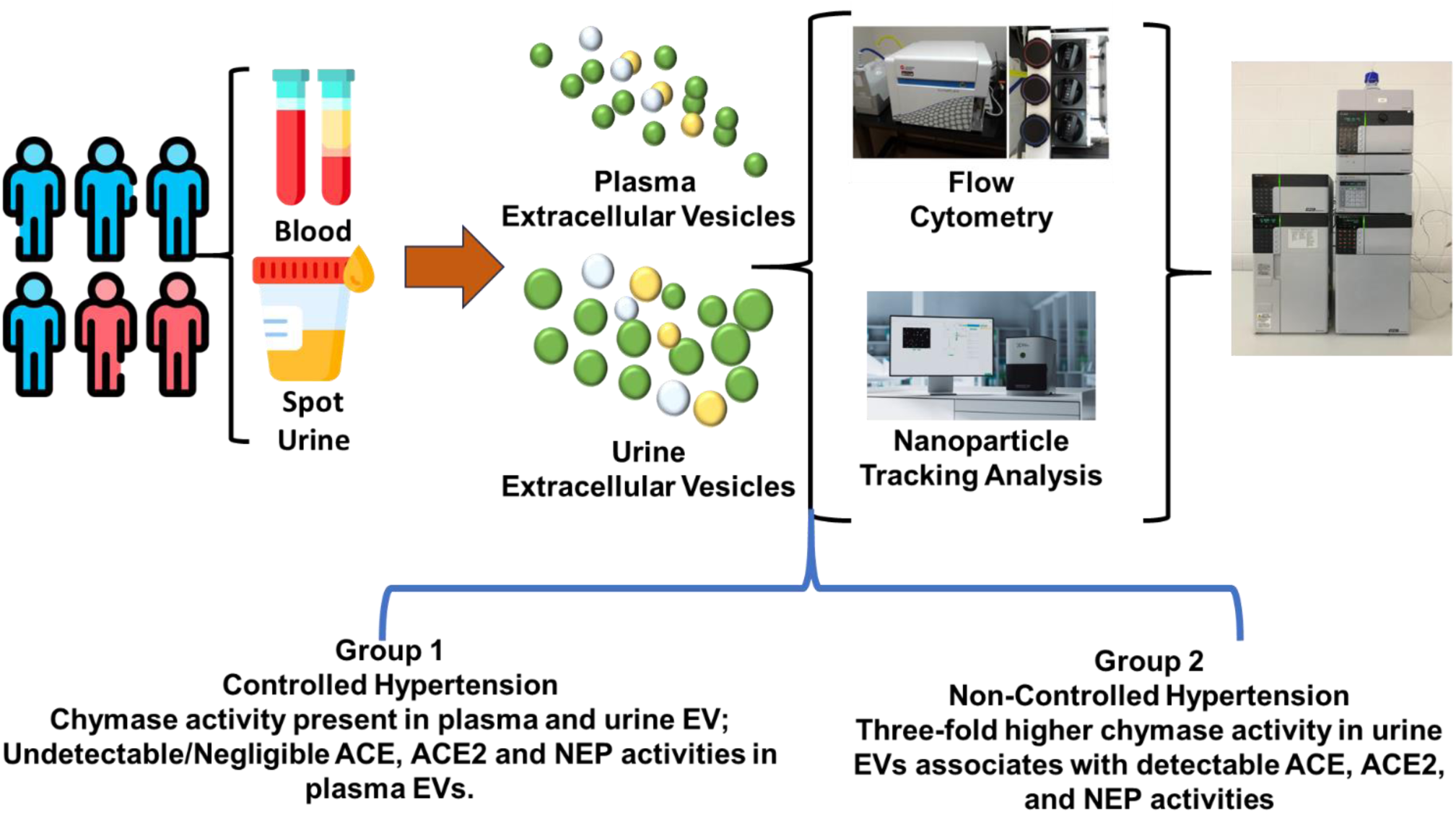

## INTRODUCTION

Precision medicine is an innovative approach for improving the diagnosis of a disease by including individual differences in people’s genes, environments, and lifestyles ^1^. A critical component of precision medicine depends on validating basic science technologies in the clinical settings. Liquid biopsies, first introduced for diagnosing cancerous cells in blood, urine, and even cerebrospinal fluid, have yielded impressive results in detecting and treating tumors ^2^. An emerging literature documents the potential of extracellular vesicles (EVs) as cardiovascular disease biomarkers and predictors of poor clinical outcomes ^3-6^. EVs are heterogeneous lipid-bound vesicles released in the extracellular space by all cell types for maintaining cellular homeostasis and cell-to-cell communication ^7^. Exosomes are endosomal-derived small extracellular vesicles (sEV), with a size less than 200 nm. These vesicles are well known to transport a variety of bioactive molecules that include proteins (e.g., cytokines, chemokines, cytoplasmic and membrane proteins, and cellular receptors), nucleic acids (e.g., long non-coding RNAs, mRNAs, and DNA), and metabolites. The cargo of these vesicles relates to the pathophysiological state of the parent cells. In support of the importance of characterizing EVs as a tissue-specific biomarker, we recently reported that EVs in the plasma could inform about corresponding molecular and metabolic alterations in the visceral adipose tissues of diet-induced obese mice ^8^. The relevance of these experimental observations was further strengthened by the parallel finding of augmented inflammation-related molecules in adipose tissue-derived Evs isolated from the blood of obese patients ^8^. Similarly, we recently reported the usefulness of tissue-specific sEV in plasma as usefulness biomarkers to assess neurocognitive decline as well as to predict treatment response in individuals with mild cognitive impairment ^9-11^. These studies support that biofluid-derived sEV could offer biomarkers for early diagnosis and identification of novel molecular targets for various pathologies.

A heightened renin angiotensin system (RAS) activity in human hypertension is acknowledged and consistent with the beneficial action of angiotensin converting enzyme (ACE) inhibitors and angiotensin II (Ang II) receptor blockers (ARBs) in blood pressure control and halting or reversing adverse cardiovascular remodeling ^12^. Although multiple factors contribute to suboptimal control of human hypertension and its cardiovascular sequela, increased attention to compensatory activation of alternate enzymatic pathways for Ang II-induced pathology following RAS pharmacotherapy is resurfacing ^13,14^. The recent identification of angiotensin-(1-12) [Ang-(1‒12)] as an alternate tissue forming Ang II substrate by chymase has provided an experimental basis for targeting non-renin and ACE-independent mechanisms for hypertension ^15-19^. The demonstration of high plasma Ang-(1‒12) levels in primary hypertensive patients naïve to antihypertensive therapy ^20^ or diagnosed as resistant hypertension ^21^ underscores the contribution of alternate Ang II forming mechanisms independent of renin and ACE ^14,22^. The importance of alternate pathways for Ang II contribution to cardiovascular disease explains the existence of a large residual risk for cardiovascular events in primary hypertensive patients treated with RAS inhibitors ^14,22^.

A vexing problem in accepting the criticality of chymase as a primary cardiac and tissue Ang II forming enzyme is that the intracellular location of the enzyme prevents it from accessing circulating or extracellularly present angiotensin I (Ang I) ^23,24^. This issue has led some investigators to exclude chymase as a tissue-residing mechanism for the catalytic conversion of Ang I into Ang II ^23,25,26^. This equivocal interpretation of RAS biotransformation mechanisms may be put to rest by our finding of chymase uptake by rat cardiomyocytes 24 h after intravenous injection of EVs isolated from the pericardial fluid of patients undergoing cardiac surgery ^27^. The present study amplifies upon our previous observations ^27^ by characterizing for the first time the presence of chymase and other angiotensins forming enzymes in plasma and urine EVs obtained from primary hypertensive patients. The results demonstrate the presence of bioactive chymase and other RAS enzymes in plasma and urinary EVs for the first time. Further, we also observed a significantly high chymase activity in urinary EVs from patients with control or uncontrolled hypertension.

## METHODS

Thirty-four adult patients (age range 31 – 80 years; 5 females) attending the Trinity Hypertension Research Institute (Carrollton, TX) for a history of hypertension and cardiovascular disease were included after consenting to participate in the study. The study, conducted according to the guidelines of good clinical practice (GCP), was approved by the Sterling Institutional Review Board, Atlanta, GA 30339 (Sterling IRB number 7175-HAPunzi).

Demographic information (age, sex, and ethnicity), clinical characteristics [body weight, body mass index (BMI), a comprehensive physical examination, and previous use of medications) were recorded in all patients (**Table 1**). Systolic and diastolic blood pressures were determined in three consecutive measures spaced two to five minutes apart with a calibrated mercury sphygmomanometer. Afterward, a venous blood sample (7 mL) was obtained from an antecubital vein. The blood was collected in a tube containing a peptidase inhibitor cocktail [1, 10-ortho-phenanthroline (0.5 mM); p-Hydroxymercuribenzoate (1 mM); pepstatin A (125 µM)], as described by us elsewhere ^28^. In addition, a 20 mL spot urine sample was obtained from all 34 patients. Blood tubes were centrifuged at 3,000 g for 10 min, and 1.0 mL aliquots of plasma and 5.0 mL of spot urine were immediately stored at -80° C until assayed.

**Table 1.** Demographics and Clinical Characteristics.

| Variable | Values |  |
| --- | --- | --- |
| | Mean ± SD | Range |
| Age, years | 63 ± 11 | 31 – 80 |
| Sex, (Male/Female) | 29/5 |  |
| Ethnicity, (Caucasian/Hispanic) | 25/9 |  |
| Body weight, Kg (n= 34) | 97 ± 24 | 64 – 179 |
| **<u>Group I. Controlled Hypertensives</u>** |  |  |
| Systolic Blood Pressure, mm Hg | 128 ± 9 | 110 – 139 |
| Diastolic Blood Pressure, mm Hg | 82 ± 9 | 69 – 99 |
| Mean Arterial Pressure, mm Hg | 97 ± 8 | 84 – 112 |
| Pulse Pressure, mm Hg | 47 ± 8 | 32 – 59 |
| Heart Rate, beats/min | 71 ± 8 | 60 – 86 |
| Body Mass Index, [$\leq 29.99$ Kg/m<sup>2</sup> (n=13)] | 27 ± 2 | 25 – 30 |
| **<u>Group II. Non-controlled Hypertensives</u>** |  |  |
| Systolic Blood Pressure, mm Hg | 147 ± 8*** | 140 – 167 |
| Diastolic Blood Pressure, mm Hg | 91 ± 9*** | 80 – 110 |
| Mean Arterial Pressure, mm Hg | 110 ± 8** | 100 – 129 |
| Pulse Pressure, mm Hg | 56 ± 6 | 50 – 69 |
| Heart Rate, beats/min | 74 ± 9 | 60 – 84 |
| Body Mass Index, [$\geq 30.0$ Kg/m<sup>2</sup> (n=21)] | 35 ± 6*** | 30 – 51 |

\*\*P < 0.0002 and \*\*\*P < 0.0001 versus Group I

### EV Isolation

We isolated EVs from plasma by a modified immunoprecipitation method utilizing ExoQuickTM (EXOQ20A-1, System Biosciences, CA, USA) as described previously ^9,10^. Briefly, 500 µl of PBS (Ca^2+^ and Mg^2+^ free) was added to 500 µl of plasma and subsequently centrifuged at 500 g for 5 min, 2,000 g for an additional 5 min, and at 10,000 g for a final 30 min at 4° C to remove the larger vesicles. The collected supernatant was mixed with 300 µl of Thromboplastin-D (176065; Fisher Scientific, Hampton, USA) and incubated for 1 hr at room temperature (RT). After incubation, 700 μL of PBS [Ca^2+^ and Mg^2+^ free and containing a cocktail of protease and phosphate inhibitors (ThermoFisher, Massachusetts, USA, Cat# 1861284)] was added and centrifuged at 1,500 g for 20 min at RT. No protease and phosphate inhibitors were added in EVs isolated for RAS activity measurements. The supernatant was then mixed with 256 μL of ExoQuickTM and incubated for an additional 1 hr at 4° C before centrifugation at 1,500 g for 30 min at 4° C. Finally, the pellet was resuspended in 200 µL of 0.1 μm filtered PBS and stored at - 80° C until analysis.

EVs from urine were collected by an ultracentrifugation method. Briefly, 5.0 mL of urine was centrifuged at 300 g for 5 minutes, 2,000 g for an additional 5 minutes, and 10,000 g for a final 30 minutes at 4° C. The urine supernatant was passed through a 0.22 µm filter and centrifuged at 100,000 g for 70 min at 4° C to pellet the EVs. Finally, the pellet was resuspended in 200 µL of 0.1 μm filtered PBS and stored at -80° C until analysis.

### Immunogold Labeling and Transmission Electron Microscopy (TEM)

The EVs were visualized using TEM after immunogold labeled as previously described by us ^10^. Briefly, the isolated EVs were fixed with 4% paraformaldehyde for 10 min at RT and adsorbed on 200 mesh Copper grids (with carbon-coated formvar film) activated with 100% ethanol. The grids were incubated at RT for 1 hr, washed 3 times (5 min each) with PBS, followed by 3 times (5 min each) with 50 mM glycine. Grids were blocked with blocking buffer (0.5% BSA in PBS) for 30 min at RT and then incubated overnight with rabbit anti-CD63 primary antibody (1:100 dilution, Abcam, Massachusetts, USA) at 4° C. The grids were then washed 3 times (5 min each) with blocking buffer (0.5% BSA in PBS) and incubated with appropriate gold-labeled secondary antibody for 2 hr at RT in the dark. The grids were washed 3 times (5 min each) with 0.1% PBST with 0.5% BSA and incubated with 2.5% glutaraldehyde for 5 min. Grids were then washed 7-timed (5 min each) with 0.1% PBST with 0.5% BSA and incubated with 1% uranyl acetate for 1 min. Finally, the grids were washed with distilled water for 2 min and imaged using TEM (FEI Tecnai Spirit transmission electron microscope system, Oregon, USA) at 98,000x magnification.

### Flow Cytometry

Flow cytometry was performed to assess the surface expression of exosomal tetraspanin markers (CD63, CD9 and CD81), as described by us previously ^8,10^. Briefly, EVs were labeled with membrane labeling dye CellBrite 488 (Biotium, California, USA) with or without the CD63-PE (353004), CD81-PE (349506) (both from BioLegend, CA, USA), or CD9-APC (Invitrogen:17-0291-82) antibodies. EVs without dye were used as a control to set the gate for positively labeled EVs. EVs labeled with dye but without CD63-PE/ CD81-PE/CD9-APC antibodies were used to set the gate for APC/ PE positive events. CD63-PE, CD81-PE, and CD9-APC antibodies and dye at the same dilution in PBS (filtered through a 0.1-micron filter) were also analyzed. All samples were acquired on CytoFlex (Beckman Coulter Life Science, Indianapolis, United States) for 60 sec at a low flow rate. Filtered PBS was run for 60 sec in between the samples.

### Nanoparticle Tracking Analysis (NTA)

The quantification of hydrodynamic size and concentration of EVs were analyzed using NTA (Nanosight NS300, Malvern Instruments, UK), equipped with violet laser (405 nm) ^10^. The instrument was prepared using PBS (pH 7.4) at 25° C temperature. Five videos of 30 seconds were recorded for every sample. The average of five videos was presented as the final size and concentration of the EVs

### Western blotting

Twenty-five μg of urine EVs were separated on the 12% SDS page gel and transferred to a nitrocellulose membrane. Blots were blocked with 5% non-fat milk for 1 hr and incubated with anti-CD63 (1:500; PA5-92370, Invitrogen), TSG 101 (1:500; ab83, Abcam), Alix (1:500, ab88388, Abcam), calnexin (1:500, ab22595, Abcam) and GM130 (1:500, NBP2-53420, Novus biologicals) antibody for overnight at 4° C. Membranes were washed with 0.1% PBST and incubated with appropriate secondary antibody for 2 hrs at RT. Blots were developed with enhanced chemiluminescence (ECL) reagent (Bio-Rad, California, USA).

### RAS enzyme activity assays in EVs

RAS enzymes activities [chymase, ACE, ACE2, and neprilysin (NEP)] were measured in isolated EVs from plasma and urine samples by HPLC using highly purified radiolabeled substrates as described by us earlier ^21^. Briefly, EVs (25-50 µg protein per 200 µL reaction mixture) were pre-incubated for 15 min in the presence or absence of chymostatin, lisinopril, and MLN-4760 ^29^. The peptidase inhibitors were diluted in an assay buffer containing 50 mM Tris-HCl + 150 mM NaCl (pH 8.0) for samples employed to determine chymase activity or 25 mM HEPES + 125 mM NaCl + 10 µM of ZnCl2 (pH 7.4) for the enzymatic detection of ACE, ACE2, and NEP activities. After pre-incubation of EVs with different combinations of inhibitor cocktail, the radiolabeled substrate [1 nmol/L each; ^125^I-Ang-(1-12) for chymase/ACE/NEP or ^125^I-Ang II for ACE2] was added to the reaction medium and incubated overnight (∼20 h) at 37° C. At the end of the incubation, the reaction was stopped by adding an equal volume of ice-cold 1% phosphoric acid. The samples were filtered through a 0.22 µm PVDF membrane syringeless filter device and were injected on the C18 column to separate the substrate and products by HPLC using the linear gradient 10% - 50% mobile phase B (80% acetonitrile/0.1% phosphoric acid) at 32° C (flow rate 0.35 mL/min), and analyzed with the Shimadzu LCSolution (Kyoto, Japan) acquisition software. Enzyme activities were calculated based on the amount of parent ^125^I-Ang substrate hydrolyzed into specific ^125^I-Ang products by the EVs in the absence and the presence of specific inhibitors. All enzyme activities are reported as fmol/min^/^mg.

### Radiolabeling of angiotensin peptides and purification

Human Ang-(1-12) and Ang II were radiolabeled with ^125^Iodine-[^125^I] at the tyrosine 4 residue using oxidant chloramine-T. Briefly, the radiolabeled substrates (^125^I-Ang-(1-12) and ^125^I-Ang II) were purified by HPLC using a linear gradient 10% - 50% mobile phase B (80% acetonitrile/0.1% phosphoric acid) at 32° C (flow rate of 0.35 mL/min). The eluted radiolabeled substrate fractions were monitored by an in-line flow-through gamma detector (BioScan Inc., Washington, DC) as well as by an in-line flow-through Shimadzu SPD UV detector (215 nm) to check the un-labeled Ang-(1-12) impurity ^21^. Products were identified by comparison of retention times of synthetic [^125^I] standard Ang-(1-12) and Ang II peptides. The highly purified radiolabeled angiotensin substrates (purity ≥99%) were used to measure enzyme activity.

### Statistical Analysis

Statistical analysis was performed using IBM SPSS Statistics version 26.0 (IBM Corporation, New York, USA) and GraphPad PRISM software (San Diego, CA). Normal data distribution was evaluated using the Shapiro-Wilk Normality and Kolmogorov-Smirnov goodness-of-fit tests. Log transformation was performed because enzyme activity values failed the Shapiro-Wilk Normality test. After log transformation, Pearson correlation tests were used to determine relationships between RAS enzyme activities and other clinical variables. We tested differences in values for log-transformed RAS enzyme activities among hypertensive groups using mixed-mode Analysis of Variance (ANOVA). The student’s t-test was used to evaluate differences between the two groups. A two-sided P < 0.05 was considered statistically significant. Values are means ± standard deviations (SD) unless stated otherwise.

## RESULTS

Demographic and clinical characteristics are documented in **Table 1**. Twenty nine of 34 patients were-male (85%); 25 reported being Caucasian (74%), and 9 declared themselves as Hispanics (26%). Thirteen out of 34 patients were classified as overweight (BMI, 27 ± 2 Kg/m^2^), while 21 others were classified as obese (BMI, 35 ± 6 Kg/m^2^) (**Table 1**). Although 85% of patients (29 of 34) included in the study were using antihypertensive medications, the blood pressure was controlled in 24 patients (Group I, **Table 1**) and not controlled in 10 others (Group II, **Table 1**) using a cut-off value of ≥140/90 mm Hg [Stage 2 hypertension ^30^]. Neither differences in risk factors nor the use of different doses and classes of antihypertensive agents influenced whether patients were included in Group I or Group II.

Plasma and urinary small EVs isolated from hypertensive patients were characterized according to the criterion defined for minimal information for studies of extracellular vesicles (MISEV, 2018) guidelines ^31^. The presence of the exosomal marker protein CD63 on the surface of EVs was validated by immunogold labeling followed by transmission electron microscopy (TEM) (**Figure 1A**). The plasma and urinary EVs isolated from the Group I (controlled hypertensive) and Group II (non-controlled hypertensive) patients were further characterized for surface markers (CD63, CD9, and CD81) by flow cytometry (**Figure 1B**, scatter plots and bar graphs). No differences in expression of the surface markers CD63, CD9, and CD81 were detected in the plasma and urinary EVs of Groups I and II (**Figure 1C**). However, the surface expression of CD81 on urinary EVs was markedly lower than plasma EVs, suggesting the heterogeneity between EVs based on the source of isolation (**Figure 1C**).

**Figure 1.**
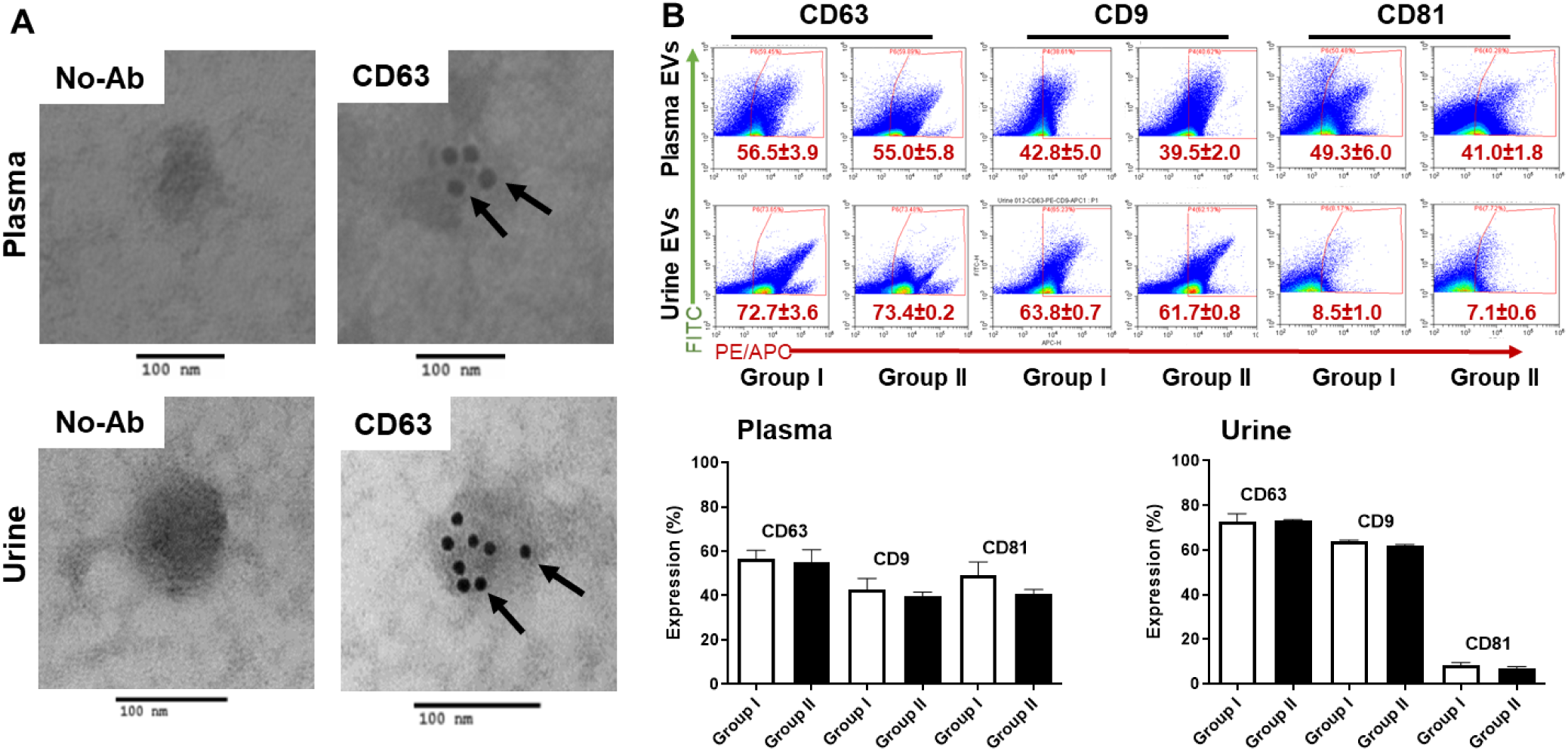
Transmission electron microscopy (TEM) and flow cytometry validate plasma and urinary EVs. The isolated plasma and urinary EVs were detected with rabbit anti-CD63 primary antibody and immuno-gold labeled anti-rabbit IgG secondary antibody by TEM (**A**), and characterized for the EVs surface markers for CD63, CD9, and CD81 by flow cytometry (**B**) as described in Methods. The flow cytometry data (scatter plots and bar graphs; Mean ± SEM) showing the expression of CD63, CD9 and CD81 on plasm and urine EVs represent three samples in each group.

Since an extensive characterization of plasma EVs had been performed by us in prior studies ^8,10,32^, we limited the current analysis to confirm the expression of exosomal biomarkers CD63, TSG101, and Alix in urinary EVs by Western blotting (**Figure 2**). Importantly, we did not observe any expression of endoplasmic reticulum protein Calnexin and Golgi protein GM130 in urinary EVs, confirming the purity of isolated EVs.

**Figure 2.**
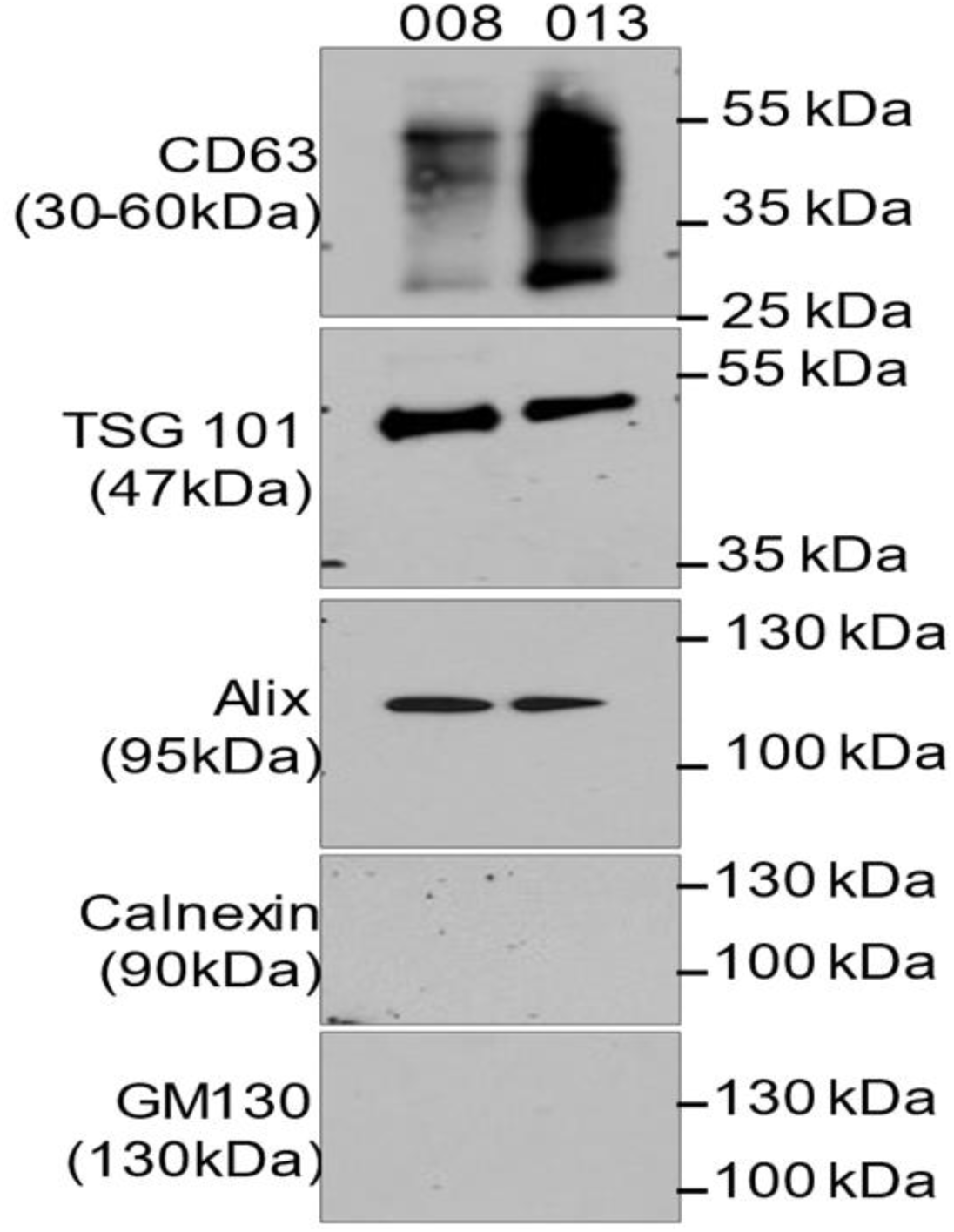
Representative immunoblots show the expression of exosome markers in urinary EVs. Urinary EVs were isolated from two samples and characterized by Western blotting for the expression of mentioned proteins.

Using NTA, the plasma and urine EVs concentration (particles/mL) and EVs percentage for various size ranges (>50 nm, 51-100 nm, 101-150 nm, 151-200 nm, 201-250 nm, 251-300 and >300 nm) were analyzed and compared between the groups in these patients (**Figure 3**). Interestingly, EVs size in urine were significantly larger in size (P < 0.0001) as compared to plasma EVs (**Figure 3A**). Further analysis based on EVs size showed that the concentration of small EVs (<200 nm) in plasma and urine were 81% and 69%, respectively. A higher concentration of medium-to-large EVs (>200 nm) were detected in urine (31%) compared to plasma (19%) (**Figure 3B and 3C**). Furthermore, the size and concentrations of plasma and urine EVs were compared between the two groups of patients. No differences in the size of plasma EVs were found between Group I and Group II patients (**Figure 3D**). No significant differences in plasma EVs concentrations were found when compared with corresponding EVs sizes between Group I (<200 nm = 80% and >200 nm = 20%) and Group II (<200 nm = 82% and >200 nm = 18%) patients (**Figure 3E and 3F**). However, urinary EV sizes were significantly larger in Group II patients (**Figure 3G**; P = 0.02 vs Group I). Whereas, in urine, a slightly higher percentage of medium-to-large EVs (Group II >200 nm = 36%) were detected when compared to Group I (>200 nm = 29%) patients (**Figure 3H and 3I**).

**Figure 3.**
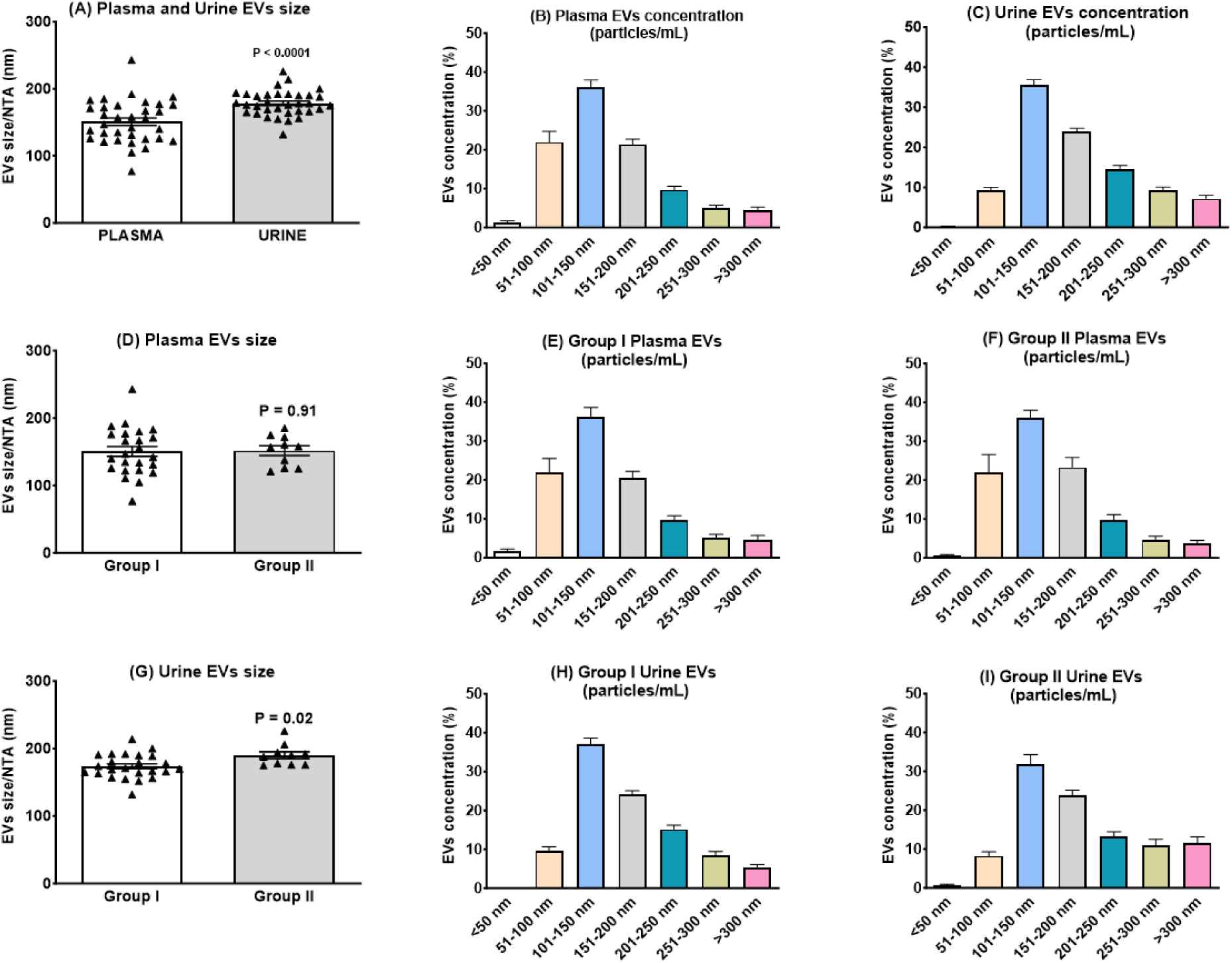
Comparative scatter graph of individual values and bars with mean ± SE of EVs average size and concentrations (particles/mL) in plasma and urine and when grouped as a function based on systolic blood pressure <140 mm Hg (Group I) or ≥140 mm Hg (Group II).

As illustrated in **Figure 4**, chymase activity in plasma and urinary EVs averaged 0.31 ± 0.18 (95% CI: 0.25 ─ 0.37) fmol/mg/min and 1.00 ± 0.63 (95% CI: 0.78 ─ 1.23) fmol/mg/min, respectively. Compared to chymase, the plasma ACE, ACE2 and NEP activities were negligible (0.01 ± 0.03, 0.01 ± 0.01 and 0.01 ± 0.03) fmol/mg/min, respectively). On the other hand, the enzymatic activities of the three enzymes were substantially greater in urinary EVs (ACE 0.26 ± 0.22, ACE2 0.07 ± 0.14 and NEP ACE2 0.87 ± 0.42) fmol/mg/min, respectively, compared to the corresponding enzyme activities expressed in plasma EVs (**Figure 4, top panel**). Urinary EVs’ chymase activity (1.00 ± 0.63 fmol/mg/min) was more than 3-fold greater than in plasma EVs. Urinary EVs also showed higher activity of NEP (0.87 ± 0.42 fmol/mg/min) (**Figure 4, bottom panel**).

**Figure 4.**
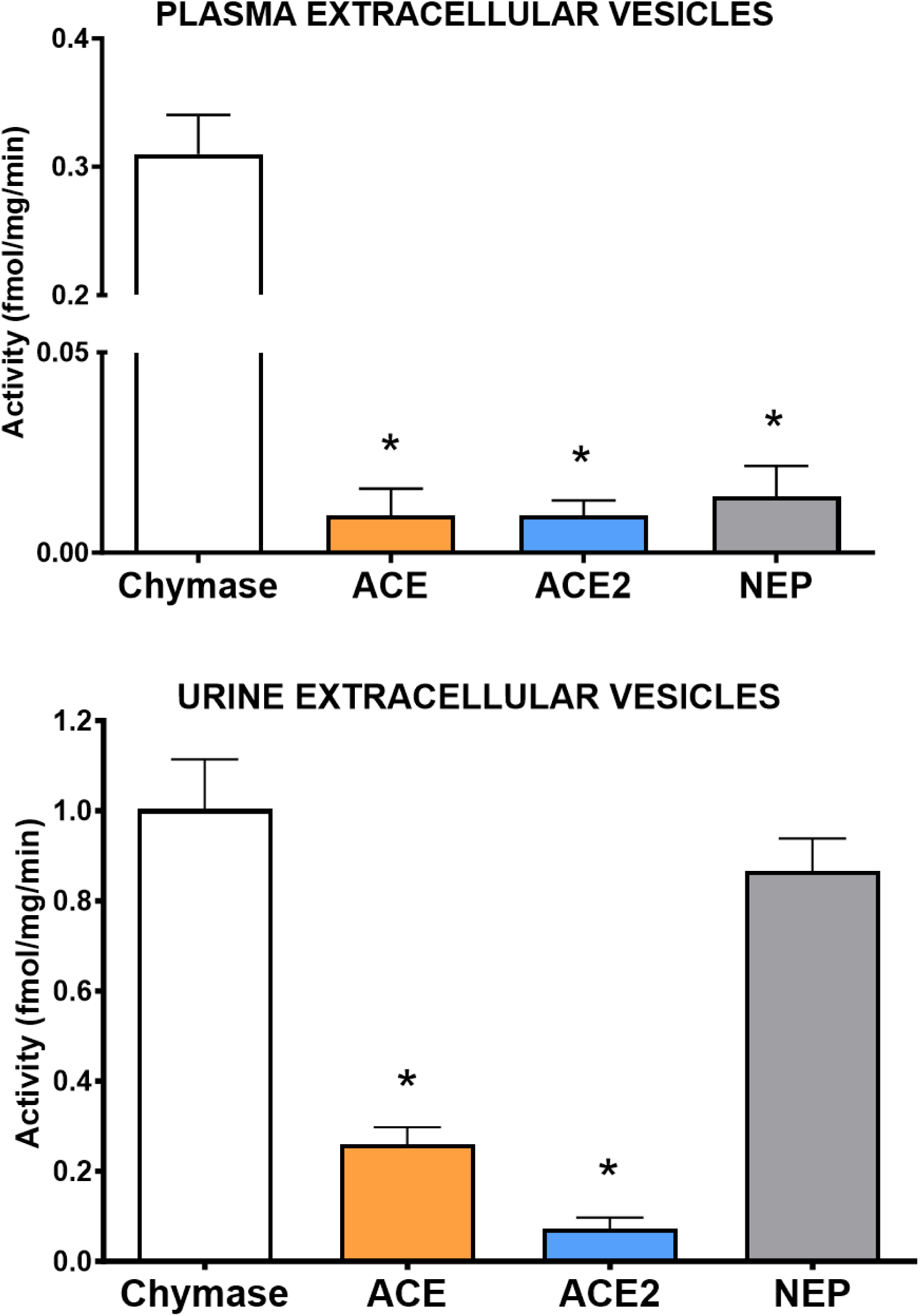
Comparative enzymatic activity of chymase, angiotensin converting enzyme (ACE), angiotensin converting enzyme 2 (ACE2), and neprilysin (NEP) in EVs isolated from the plasma (top graph) and urine (bottom graph) of hypertensive patients. Values are means ± SD. *P < 0.05 versus chymase activity.

Differences in the enzymatic activities of plasma and urinary EVs were further compared in patients within Group I (n=21) and Group II (n=8). Plasma and urinary EVs (**Figure 5A and 5B**) showed higher values of chymase activity in patients with systolic blood pressure (SBP) ≥140 mm Hg [0.44 ± 0.16 (95% CI: 0.26 ─ 0.38) fmol/mg/min as compared to SBP <140 mm Hg [0.32 ± 0.13 (95% CI: 0.26 ─ 0.38) fmol/mg/min]. Furthermore, the size of the EVs isolated from the urine was significantly larger compared to plasma EVs in Group II patients (**Figure 5A**). No differences in plasma EVs size were found between patients included in the two groups (**Figure 5B**). However, urinary EVs were significantly larger in Group II patients (**Figure 5C**; Mean difference change: 16.52 ± 6.584 nm, P = 0.02). As shown in **Figure 5B-D**, the ACE, ACE2, and NEP activities in plasma EVs were negligible/undetectable in both groups of patients. Due to negligible/undetectable values in most plasma EVs, the enzymatic activities were measured only in 7-8 samples in each group. No differences were found in the urinary EVs ACE, ACE2, and NEP enzyme activities in urinary EVs between Group I and Group II (**Figure 5F-H**).

**Figure 5.**
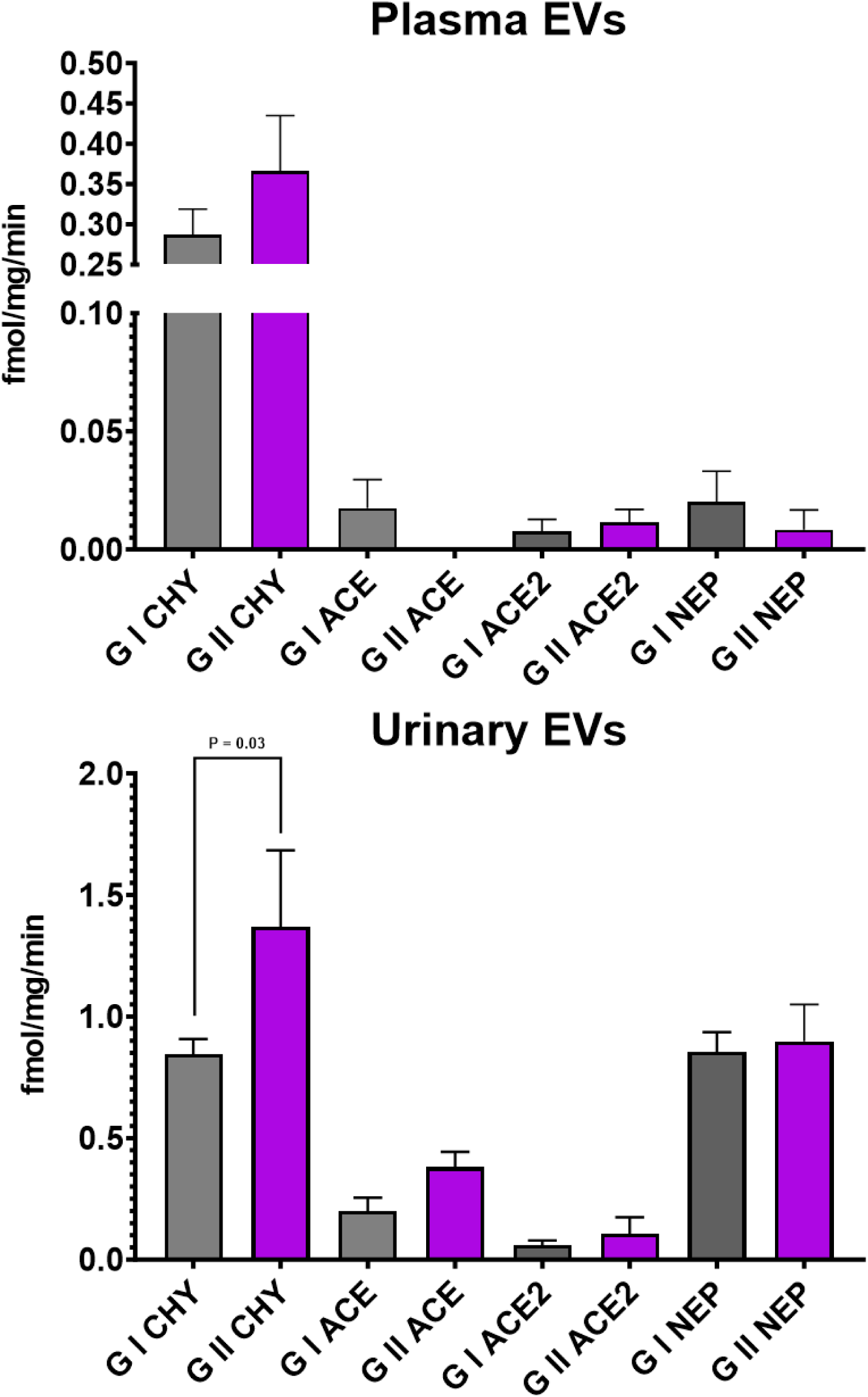
RAS enzyme activity in plasma and urinary Evs from controlled (G I) and not controlled (G II) hypertensive patients. Values are means ± SE.

A pooled multiple correlation analysis of enzyme activities from EVs isolated from plasma and urine of controlled and not-controlled hypertensive patients is documented in **Table 2**. Plasma EVs chymase activity was strongly correlated with both urinary EVs chymase (P = 0.003) and NEP (P = 0.05) hydrolytic activities (**Table 2**). Urinary EVs chymase activity was correlated with urinary EVs ACE (P = 0.0001), ACE2 (P = 0.005), and NEP (P = 0.0001) activity (**Table 2**), while urinary EVs ACE was highly correlated with both urinary EVs ACE2 (P = 0.0001) and NEP (P = 0.0001) activities. Further, urinary EVs ACE2 was highly correlated with urinary EVs NEP (P = 0.0001) activity (**Table 2**). No significant differences were observed for other variables (including urine and plasma EVs sizes).

**TABLE 2.** Multiple Correlation Analysis.

| **Variables** | **Plasma Chymase**<br>**(fmol/mg/min)** | **Urine Chymase**<br>**(fmol/mg/min)** | **Urine ACE**<br>**(fmol/mg/min)** | **Urine ACE2**<br>**(fmol/mg/min)** | **Urine Neprilysin**<br>**(fmol/mg/min)** |
| --- | --- | --- | --- | --- | --- |
| **Plasma Chymase,**<br>**(fmol/mg/min)** | **1.00** | 0.47<br>(P=0.003) | 0.23<br>(n.s.) | 0.02<br>(n.s.) | 0.31<br>(P=0.05) |
| **Urine Chymase,**<br>**(fmol/mg/min)** |  | **1.00** | 0.66<br>(P < 0.0001) | 0.34<br>(P=0.05) | 0.59<br>(P < 0.0001) |
| **Urine ACE,**<br>**(fmol/mg/min)** |  |  | **1.00** | 0.53<br>(P < 0.0001) | 0.71<br>(P < 0.0001) |
| **Urine ACE2,**<br>**(fmol/mg/min)** |  |  |  | **1.00** | 0.64<br>(P < 0.0001) |

(n.s), not significant (P > 0.05)

## DISCUSSION

EVs have emerged as key players in local and distant inter-cellular communication in healthy and disease conditions. We show for the first time that well-characterized EVs from the blood and urine of hypertensive patients express chymase activity in proportions that in plasma are approximately 3-fold higher than the corresponding hydrolytic activities of ACE, ACE2, and even NEP. We further show that EVs isolated from the urine of hypertensive subjects are significantly larger in size and exhibit higher values of chymase, ACE, and ACE2 activities. In addition, we demonstrate that NEP activity in urinary EVs is not different than the chymase activity. The potential of these EVs’ enzymatic activities as biomarkers of RAS activity in hypertensive individuals is further strengthened by the correlative demonstration of higher chymase activity in EVs isolated from both the blood and urine of medicated but not controlled hypertensive patients. The data reported in this study establishes a foundation for understanding how circulating blood and urinary EVs act as a dynamic vehicle for transporting chymase to other organs such as the heart and kidneys. The significance of these findings is buttressed by the demonstration of increased EVs chymase cargo in hypertensive patients who are medicated but yet remain not controlled.

Human blood and urine EVs were isolated and characterized with a well-documented rigorous protocol previously published by us elsewhere ^9,10^. The array employed in these studies included highly specific exosomal marker proteins ^33,34^. In our study, urinary EVs were bigger than those isolated from the plasma. The size of both plasma and urinary EVs is within the size of exosomal vesicles reported in the literature ^35^.

A budding literature identifies exosomes as vehicles for transferring ACE to vascular smooth muscle cells in SHR ^36^. As reviewed by Ren and Zhang ^37^, microRNAs isolated from the blood of genetically hypertensive rats can exert significant structural changes in the vasculature, reduce oxidative stress, and even Mas-receptor expression. In addition, we showed recently that chymase containing EVs isolated from the pericardial fluid of patients undergoing cardiac surgery delivered human chymase to endothelial cells and the rat cardiac interstitium within 4 h post-injection ^27^.

We previously reported the presence of higher levels of circulating Ang-(1‒12) in primary hypertensive patients naïve ^20^ and not naïve ^21^ to antihypertensive therapy. The present study expands those earlier findings by concomitant demonstrating higher chymase activity in blood and urinary EVs from those treated but not controlled subjects. The demonstration of a selective increase in chymase activity in blood and urine EVs strengthens the possibility that EVs enriched with chymase activity, acting as a “liquid biopsy,” ^8^ provides insights into the evolution of the hypertensive process, the degree and location of predominant target organ damage, and the response to therapy. This interpretation is underscored by studies reporting an active role of proteins and non-coding microRNAs in circulating exosomes in mice with induced Ang II target organ damage ^38^, experimental atherosclerosis ^39^, cardiac fibrosis ^40^, T-cell derived EVs ^41^, hypertensive patients treated with Ang II receptor blockers (ARBs) ^42^, EVs harvested from the pericardial fluid of patients undergoing open heart surgery ^43,44^, and experimental models of chronic kidney disease ^45,46^.

EVs mediate cell-to-cell communication via the transfer of their cargo to the recipient cells. This is the first demonstration of RAS enzyme activity in blood and urine EVs from hypertensive patients. This new finding suggests selective biogenesis of EVs expressing RAS enzymes in the kidney or the extrusion of selective cargo from cell membranes, as reported for the transport of microRNAs ^47^. The lower chymase activity in blood EVs is associated with traces of enzymatic hydrolytic activity for ACE, ACE2, and NEP. This finding suggests a selective specificity of the cargo present in these EVs. It may be argued that endogenously circulating serine chymase inhibitors may explain the low chymase enzymatic activity demonstrated in blood EVs ^48^. This is unlikely since circulating serpins would have had no access to the chymase residing inside the EVs ^49^. In addition, detecting only trace ACE, ACE2, and NEP enzymatic activities in the circulating EVs suggests a preferential selection of the chymase cargo and, indirectly, excludes the potential role of endogenous inhibitors from interfering with the assay’s specificity. The higher values of RAS activities in urinary EVs could be associated with the larger size of the EVs. While this finding is in keeping with the observation that EVs size correlates with higher cargo levels ^35^, it certainly needs further investigation.

The pathogenic importance of chymase as contributing to cardiovascular pathology remains controversial, in part because its primary intracellular location has been equivocally interpreted to exclude the enzyme from contact with interstitial or circulating angiotensinogen or angiotensin I ^23,50^. While Danser and colleagues ^23,51^ persist in dismissing chymase as a contributor to tissue Ang II formation and cardio-renal dysfunction, the data reported here provides a novel mechanism by which chymase, residing in the protected environment of EV, could transport the enzyme to cells in cardiovascular tissues. Transferring proteins and lipids locally and systemically to other organs is a fundamental function of EVs ^35^. Our earlier demonstration of human EV chymase incorporation into rat’s cardiomyocytes evidence that circulating exosomes may constitute a vehicle for the transport of chymase into organs that express angiotensinogen and Ang-(1‒12) such as the heart, the kidneys, and even the thymus ^13,18,52-54^.

Chymase is abundant in mast cells, contributing to no less than 25% of the total protein content ^55^. As an ACE-independent source of Ang II formation, mast cell chymase plays a crucial role in the development of kidney disease, chronic inflammation, and kidney graft rejection as evidenced by increased mast cell accumulation in renal tubules and the adventitia of intrarenal vessels ^56^. Immunohistochemical analysis of human renal biopsy specimens visualizes an increased number of mast cells infiltrating distal renal tubules preferentially or in contact with fibroblasts, lymphocytes, or macrophages in the cortical renal interstitium ^57^. Ang II production by chymase appears to be a major source of renal Ang II production in kidneys after a high salt intake and diabetic nephropathy ^55^. Chymase catalytic activity to generate Ang II from Ang I is 20-fold higher than ACE ^58^. In addition, we showed that chymase activity was 25-fold higher than the ACE activity in human left atrial appendage tissues obtained from patients undergoing cardiac surgery ^15^. Although NEP has been reported to be the major Ang-(1-7)-forming enzyme in the kidney ^59,60^, chymase, not ACE, assumes a primary hydrolytic role in renal Ang II pathology and its progression to chronic kidney disease ^61^. While the origin of the enriched chymase and NEP cargo in urinary EVs remains to be established, our findings suggest a critical role of renal chymase as a significant biomarker of uncontrolled high blood pressure.

In summary, we identified chymase as a preferential cargo in circulating and urine EVs in treated hypertensive patients. This demonstration places this enzyme as a major signaling vector in the processes accounting for cardiovascular pathology, chronic inflammation, and renal dysfunction in primary hypertension. In addition, we postulate that chymase activity in EVs is a potential biomarker of the ability of antihypertensive medications to interfere with the disease process.

## Data Availability

Corresponding author/co-authors ensure that the data remains widely available to the research community.

## Acknowledgments

The data reported in this study was funded by Program Project Grant HL-051952 from the National Heart, Lung, and Blood Institute (NHLBI) (CM Ferrario) and the Institute on Aging (1 R21 AG070371-01) (CM Ferrario) from the National Institutes of Health. This work was supported by NIA RF1AG068629 and R01 AG061805 (GD). Wake Forest Baptist Comprehensive Cancer Center Cellular Imaging Shared Resource is supported by NCI (P30CA012197, PI: Dr. Ruben A. Mesa).

## Availability of data

The data underlying this article are available in the article and in its online supplementary material.

